# Rapid increase in SARS-CoV-2 seroprevalence during the emergence of Omicron variant, Finland

**DOI:** 10.1101/2022.03.25.22272952

**Authors:** MJ Ahava, H Jarva, AJ Jääskeläinen, M Lappalainen, O Vapalahti, S Kurkela

**Affiliations:** HUS Diagnostic Center, HUSLAB, Clinical Microbiology, University of Helsinki and Helsinki University Hospital, Finland; Translational Immunology Research Program and Department of Bacteriology and Immunology, University of Helsinki, Helsinki, Finland; Department of Virology, Faculty of Medicine, University of Helsinki, Helsinki, Finland; Department of Veterinary Biosciences, Faculty of Veterinary Medicine, University of Helsinki, Helsinki, Finland

**Author notes:** **Corresponding author:** Maarit Ahava, **Email:**, **Address:** Helsinki University Hospital, HUSLAB, Virology and Immunology, P.O.B. 720 (Topeliuksenkatu 32), FIN-00029 HUS, Finland.

**Keywords:** COVID-19, SARS-CoV-2, serology, surveillance

## Abstract

**Objectives:** The aim of this study was to assess changes in exposure and prevalence of SARS-CoV-2 infection during the first months of emergence of Omicron variant in the Greater Helsinki area, Finland.

**Methods:** A prospective seroepidemiological survey of SARS-CoV-2 was conducted on 1,600 serum specimens sent to Helsinki University Hospital Laboratory (HUSLAB) for HIV serology between 15 November 2021 and 6 March 2022 (calendar weeks 46/2021 – 9/2022). For each calendar week, 100 serum specimens were randomly selected and analysed for SARS-CoV-2 IgG antibodies against nucleocapsid (N) and spike 1 (S1) protein with Abbott SARS-CoV-2 IgG (N protein) and SARS-CoV-2 IgG II Quant (S protein) tests, respectively.

**Results:** The prevalence of N antibodies increased from 5.2% (weeks 46-50/2021) to 28.2% (weeks 5-9/2022) during the study period. The proportion of seronegative samples as well as anti-N negative, anti-S1 positive samples decreased correspondingly from 11.6% to 3.8%, and 84.2% to 68.2%, respectively. Anti-N positive samples that were anti-S1 negative only began to appear as of week 2/2022.

**Conclusions:** A rapid increase in the N antibody prevalence was observed over the study period, suggesting a high transmission rate. A substantial proportion of COVID-19 cases remained undiagnosed during the emergence of Omicron variant in the Greater Helsinki Area, Finland.

---

The emergence of SARS-CoV-2 Omicron variant (B.1.1.529) as of November 2021 changed the epidemiology of COVID-19 with rapid upsurge of cases globally [1,2]. In Finland, the first patient case with Omicron variant was detected on 29 November 2021 [3]. In this study, we conducted a prospective seroepidemiological survey of SARS-CoV-2 in November 2021 – March 2022 in the Greater Helsinki area, Finland. Our aim was to assess changes in exposure and prevalence of SARS-CoV-2 infection during the first months of emergence of Omicron variant.

The study was institutionally approved (HUS/56/2021). Altogether 1,600 serum specimens were analyzed with Abbott SARS-CoV-2 IgG II Quant (IgG antibodies to receptor binding domain (RBD) of the S1 subunit of the S protein of SARS-CoV-2) and N antibodies with Abbott SARS-CoV-2 IgG (IgG antibodies to N protein of SARS-CoV-2) on the Alinity i analyzer. The sampling scheme included 100 specimens each week between weeks 46/2021 and 9/2022, from routine samples sent to HUS Diagnostic Center, Helsinki. The sampling frame comprised 17,000 serum specimens that were tested negative for HIV Ag/Ab between 15 November 2021 and 6 March 2022 and stored according to date of specimen. To select samples for each calendar week, a random starting point was chosen, and specimens were systematically selected until 100 specimens plus 5 spare samples were reached: the chosen 100 specimens were analyzed for SARS-CoV-2 antibodies. If the analysis failed or the specimen volume was not adequate, the sample was replaced by one of the spare samples of that calendar week.

The subgroups identified according to serostatus were I) anti-N negative, anti-S1 negative: no serological evidence of vaccine immunization or previous infection; II) anti-N negative, anti-S1 positive: seroresponse to vaccine immunization, no evidence of recent infection; III) anti-N positive, anti-S1 positive: consistent with previous infection, vaccine immunization status unknown; IV) anti-N positive, anti-S1 negative: recent infection possible, no evidence of vaccine immunization. The proportion of these subgroups was determined for each calendar week and statistical analyses were performed using IBM SPSS statistical program package, version 25., visualization was done with GraphPad Prism 8.0.1.

The study subjects’ age ranged from 11 months to 94 years (median 33 years; IQR 26-46 years), and the proportion of women was 55.2%. The baseline prevalence of N antibodies in the first five weeks of the study period (46-50/2021) was 5.2%, while in the final five weeks (5-9/2022) it was 28.2%. The proportion of seronegative samples for the corresponding time frames was 11.6% and 3.8%, and for anti-N negative, anti-S1 positive samples 84.2% and 68.2%. Figure 1 depicts the moving average of the N antibody seroprevalence over the study period: the sharpest increase was observed in those aged <30 years. In late 2021, the seroprevalence of N antibodies was consistently well below 10% but began a rapid incline as of week 1/2022 and surpassed 20% on week 3/2022.

**Figure 1:**
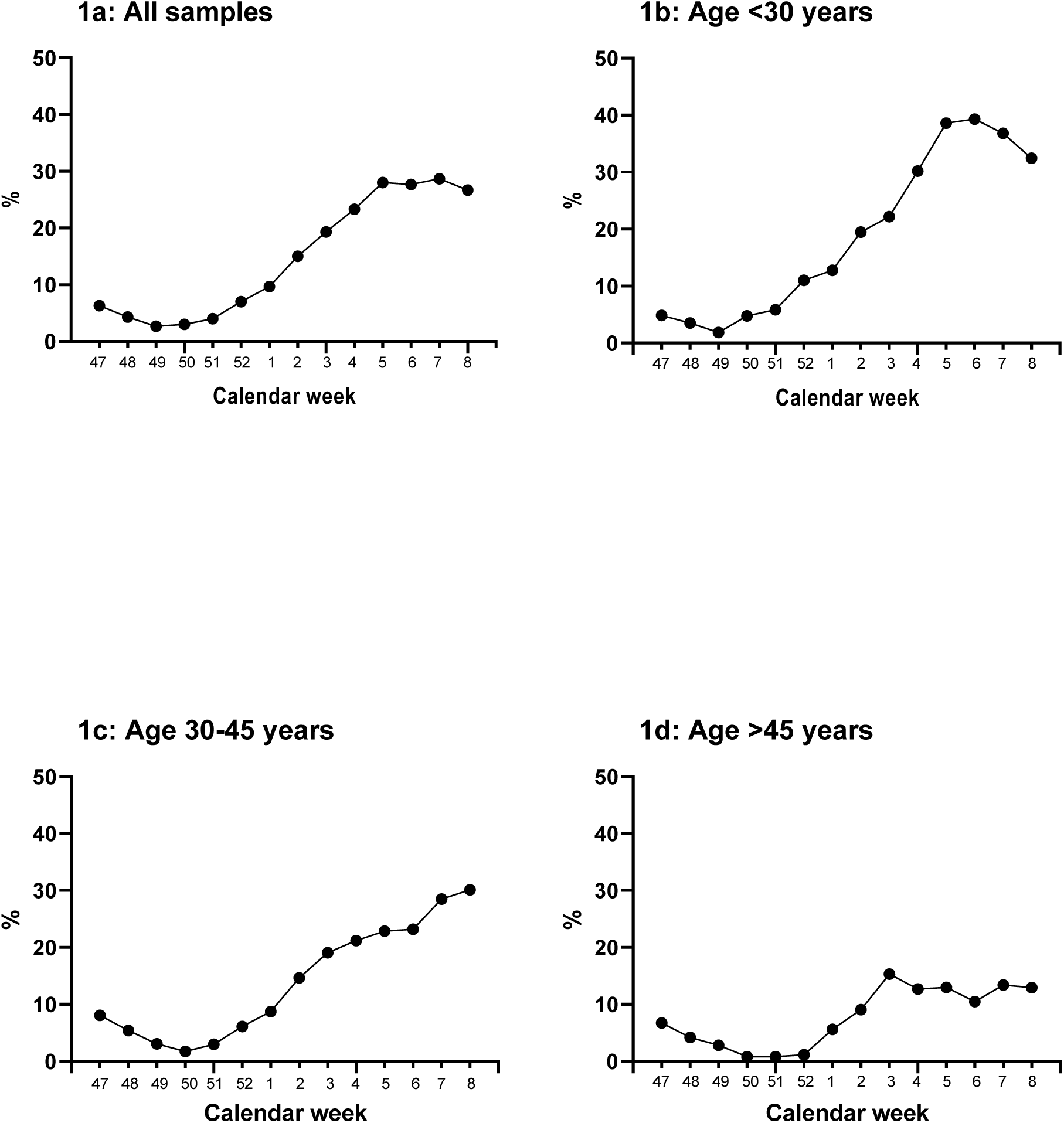
Proportion of N antibody positive samples, three-week moving average. 1a: All samples from all age groups, N=1600, 1b: Study subjects under 30 years, N=616 1c: study subjects 30-45 years, N=580, 1d: study subjects over 45 years, N=404.

Anti-N positive samples that were anti-S1 negative began to appear on week 2/2022 and represented 0.9% (14/1600) of all analyzed samples, which may reflect a diminished or delayed seroresponse against S1 during Omicron infection. The increase in anti-N positive samples (groups III and IV) was reflected as a decreasing proportion of seronegative samples (group I) towards the end of the study period. The proportions of subgroups (I-IV) per calendar week are presented in Figure 2.

**Figure 2:**
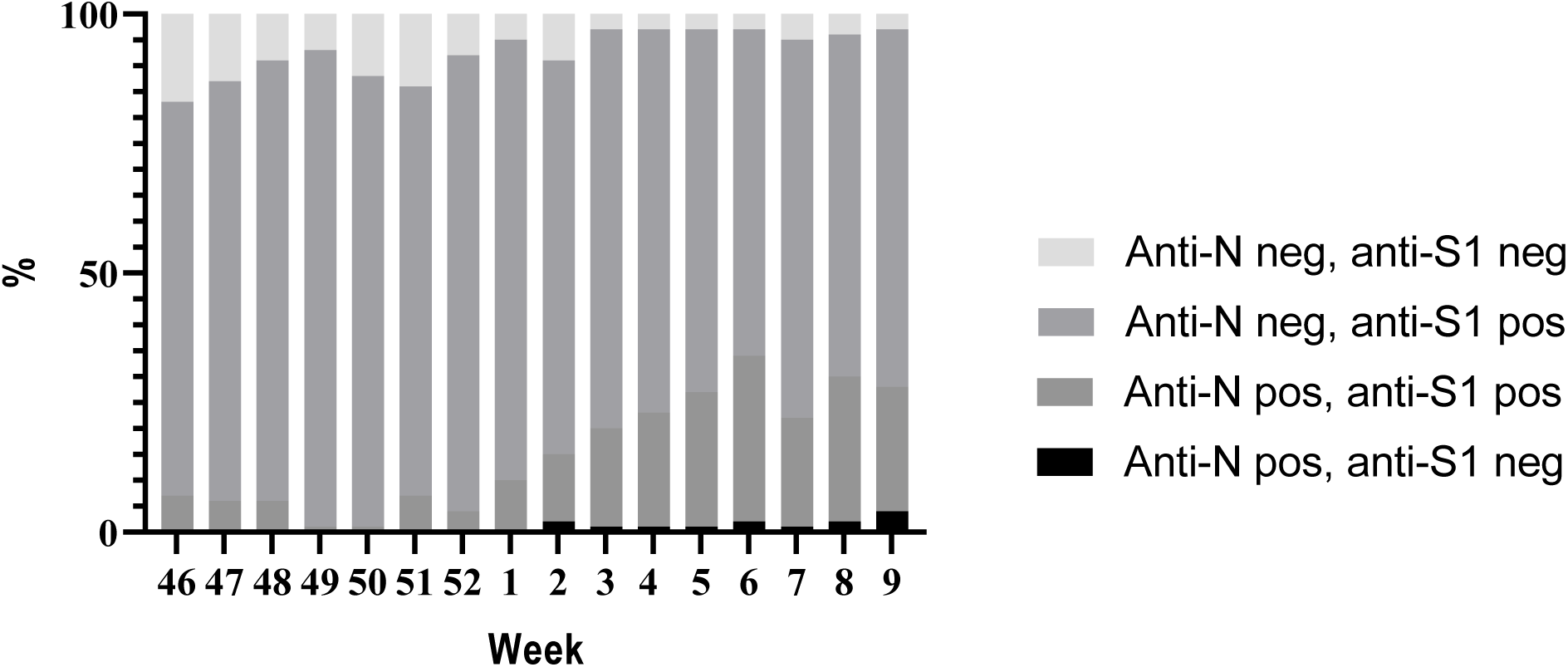
Proportions of subgroups over the study period. The increase in N antibody positive samples was reflected as a decreased proportion of both the group of completely seronegative samples and samples of immunized individuals.

By mid-December 2021, Omicron had become the primary variant in the Greater Helsinki area [3]. Soon after, our data show a rapid increase in the population level exposure to SARS-CoV-2. This indicates a high transmission rate and is in line with previous reports from elsewhere [4,5,6]. At the end of the study period (week 9/2022), 4% were seronegative in N and S antibody testing. Altogether 78% (1241/1600) had S antibodies without N antibodies, suggesting vaccine immunization without recent COVID-19 infection.

Our study design did not allow differentiation between those who had undergone COVID-19 with or without prior vaccine immunization. Also, protective immunity against SARS-CoV-2 cannot be determined by detection of antibodies to N or S antigen by enzyme immunoassays. As the purpose of this study was to provide real-time data, conducting neutralization assays was not feasible.

The present study showed a rapid increase in the N antibody prevalence, indicating that approximately 23% (comparing the first and last five weeks of the study period) of the tested individuals had contracted SARS-CoV-2 infection during the emergence of Omicron variant. While our sampling frame does not perfectly reflect the general population, the data suggest that during the study period, well beyond 300,000 individuals in the Greater Helsinki area (population 1.5 million inhabitants) underwent COVID-19, while at the same time approximately 230,000 COVID-19 cases were officially diagnosed in Greater Helsinki [7]. The present study suggests that a substantial proportion of COVID-19 cases remained undiagnosed during the emergence of Omicron, probably due to subclinical infections and diminished RT-PCR testing.

## Data Availability

The datasets generated during and/or analysed during the current study are available upon reasonable request from the corresponding author.

## Acknowledgements

We would like to thank Ms. Irina Leino and Ms. Merja Heikkinen (Department of Virology and Immunology, HUSLAB) for excellent technical assistance.

## Contribution

MA: Maarit Ahava; HJ: Hanna Jarva; AJ: Annemarjut J Jääskeläinen; ML: Maija Lappalainen; OV: Olli Vapalahti; SK: Satu Kurkela

Conceptualisation: HJ, OV, SK. Data curation: MA, HJ, SK. Formal analysis: MA, HJ, SK. Investigation: MA, HJ, AJ, ML, OV, SK. Methodology: MA, AJ, SK. Project administration: HJ, SK. Resources: ML. Validation: MA, HJ, AJ, ML, OV, SK. Writing - original draft: MA, SK. Writing - review & editing, MA, HJ, AJ, ML, OV, SK. All authors approved the final version of the manuscript.

## Data availability

The datasets generated during and/or analysed during the current study are available from the corresponding author.

## References

1. ECDC. Assessment of the further emergence and potential impact of the SARS-CoV-2 Omicron variant of concern in the context of ongoing transmission of the Delta variant of concern in the EU/EEA, 18th update. Published online 2021.

2. WHO Coronavirus (COVID-19) Dashboard. Accessed March 23, 2022. https://covid19.who.int/

3. Vauhkonen H, Truong P, Kant R et al. Introduction and rapid spread of SARS-CoV-2 Omicron variant and the dynamics of its sub-lineages BA.1 and BA.1.1, December 2021, Finland, 23 March 2022, PREPRINT (Version 1) available at Research Square. doi: 10.21203/rs.3.rs-1480433/v1

4. Baker JM, Nakayama JY, O’Hegarty M, et al. SARS-CoV-2 B.1.1.529 (Omicron) Variant Transmission Within Households — Four U.S. Jurisdictions, November 2021–February 2022. MMWR Morb Mortal Wkly Rep. 2022;71(9):341–346. doi:10.15585/MMWR.MM7109E11.

5. Brandal LT, MacDonald E, Veneti L, et al. Outbreak caused by the SARS-CoV-2 Omicron variant in Norway, November to December 2021. Eurosurveillance. 2021;26(50):2101147. doi:10.2807/1560-7917.ES.2021.26.50.2101147

6. Song JS, Lee J, Kim M, et al. Serial Intervals and Household Transmission of SARS-CoV-2 Omicron Variant, South Korea, 2021 - Volume 28, Number 3— March 2022 - Emerging Infectious Diseases journal - CDC. Emerg Infect Dis. 2022;28(3):756–759. doi:10.3201/EID2803.212607

7. THL, National Institute for Health and Welfare : National infectious disease register. Accessed March 23, 2022. https://sampo.thl.fi/pivot/prod/fi/epirapo/covid19case/fact_epirapo_covid19case;jsessionid=6A49E7FCD1DCCEB303F35730221824F1.apps5?row=dateweek20200101-509030&column=measure-444833.445356.492118.&fo=1&filter=hcdmunicipality2020-445193

